# COVID-19-induced acute respiratory failure – an exacerbation of organ-specific autoimmunity?

**DOI:** 10.1101/2020.04.27.20077180

**Authors:** D Gagiannis, J Steinestel, C Hackenbroch, M Hannemann, V Umathum, N Gebauer, M Stahl, H Witte, K Steinestel

**Affiliations:** Department of Pulmonology, Bundeswehrkrankenhaus Ulm, 89081 Ulm, Oberer Eselsberg 40, Germany; Clinic of Urology, University Hospital Augsburg, 86156 Augsburg, Stenglinstraße 2, Germany; Department of Radiology, Bundeswehrkrankenhaus Ulm, 89081 Ulm, Oberer Eselsberg 40, Germany; Department of Laboratory Medicine, Bundeswehrkrankenhaus Ulm, 89081 Ulm, Oberer Eselsberg 40, Germany; Institute of Pathology and Molecular Pathology, Bundeswehrkrankenhaus Ulm, 89081 Ulm, Oberer Eselsberg 40, Germany; Department of Hematology and Oncology, University Hospital Schleswig-Holstein Campus Luebeck, 23552 Luebeck, Ratzeburger Allee 160, Germany; Department of Hematology and Oncology, Bundeswehrkrankenhaus Ulm, 89081 Ulm, Oberer Eselsberg 40, Germany

**Author notes:** Shared senior authorship (H Witte, K Steinestel).

## Abstract

**Background:** Understanding the pathophysiology of respiratory failure (ARDS) in coronavirus disease 2019 (COVID-19) patients is of utmost importance for the development of therapeutic strategies and identification of risk factors. Since we observed clinical and histopathological similarities between COVID-19 and lung manifestations of connective tissue disease (CTD-ILD) in our clinical practice, aim of the present study is to analyze a possible role of autoimmunity in SARS-CoV-2-associated respiratory failure.

**Methods:** In this prospective, single-center trial, we enrolled 22 consecutive patients with RT-PCR-confirmed SARS-CoV-2 infection hospitalized in March and April, 2020. We performed high-resolution computed tomography (HR-CT) and full laboratory testing including autoantibody (AAB) screening (anti-ANA, SS-B/La, Scl-70, Jo-1, CENP-B, PM-Scl). Transbronchial biopsies as well as post mortem tissue samples were obtained from 3 and 2 cases, respectively, and subsequent histopathologic analysis with special emphasis on characterization of interstitial lung disease was performed.

**Results:** Twelve of 22 patients (54.5%) were male and median age was 69.0 (range: 28-88). 11 (50.0%) patients had to be undergo intensive care unit (ICU) treatment. Intubation with ventilation was required in 10/22 cases (46%). Median follow-up was 26 days. Clinical and serological parameters were comparable to previous reports. Radiological and histopathological findings were highly heterogeneous including patterns reminiscent of CTD-ILD. AAB titers ≥1:100 were detected in 10/11 (91.9%) COVID-19 patients who required ICU treatment, but in 4/11 (36.4%) patients with mild clinical course (p=0.024). Patients with AABs tended to require invasive ventilation and showed significantly more severe complications (64.3% vs. 12.5%, p=0.031). Overall COVID-19-related mortality was 18.2% among hospitalized patients at our institution.

**Conclusion:** Our findings point out serological, radiological and histomorphological similarities between COVID-19-associated ARDS and acute exacerbation of CTD-ILD. While the exact mechanism is still unknown, we postulate that SARS-CoV-2 infection might trigger or simulate a form of organ-specific autoimmunity in predisposed patients. The detection of autoantibodies might identify patients who profit from immunosuppressive therapy to prevent the development of respiratory failure.

## INTRODUCTION

Coronavirus disease 2019 (COVID-19), caused by the severe acute respiratory syndrome coronavirus 2 (SARS-CoV-2), has caused or contributed to ten thousands of deaths and led to almost complete shutdown of social and economic life in many countries [1]. Clinically, most patients with COVID-19 present with fever, dry cough and expectoration. Lymphopenia, high levels of neutrophils and high serum levels of lactate dehydrogenase, bilirubin and D-dimers have previously been identified as predictors of poor outcome [2]. Based on what is currently known about the epidemiology, COVID-19 is associated with a mortality rate between 1% and 7% [3]. The major cause of death in COVID-19 infection is acute respiratory failure (acute respiratory distress syndrome, ARDS), however, the exact mechanism how COVID-19 leads to ARDS is unclear. In the few reported cases in which tissue samples could be obtained, the authors describe diffuse alveolar damage (DAD) with an early edematous phase followed by hyaline membrane formation, desquamation of pneumocytes and an increased interstitial mononuclear infiltrate [4]. In one case, Tian et al. report loose intra-alveolar fibromyxoid proliferation reminiscent of organizing pneumonia (OP) [5]. Such combined histological patterns of ARDS and OP are observed in acute fibrinous organizing pneumonia (AFOP) and are commonly seen as rather unspecific common final paths from a variety of noxogenic stimuli that can affect the lung. These include post-infective change, trauma, burns and acute exacerbation of connective tissue disease (CTD-ILD) [6]. However, the AFOP pattern, where plug-like alveolar fibrin deposition is accompanied by alveolar and/or septal fibroblast proliferation, is accordingly observed in CTDs such as systemic lupus erythematosus (SLE), dermatomyositis and progressive systemic sclerosis (PSS) [7-9].

Previous studies have demonstrated that coronaviruses activate toll-like receptors (TLRs) on antigen-presenting cells (APCs), followed by a release of IFNα and functional exhaustion of cytotoxic T-lymphocytes (CTLs) [10,11]. These mechanisms are highly similar to development and progression of CTDs [12]. For example, functional exhaustion of CTLs is observed in SLE patients [13]. CTDs are in most cases accompanied by presence of autoantibodies (AABs): antinuclear antibodies (ANAs), antinucleolar antibodies (PM-Scl) and antibodies against host epitopes which are mostly summarized under the term extractable nuclear antibodies (ENAs): anticentromer antibodies (CENP-B), SS-B/La, Jo-1 and Scl-70 [14]. It has to be noted that these AABs can also be detected in healthy individuals and their presence is thus not entirely specific for diagnosis of CTDs [14]. However a screening dilution of 1:160 represents a cutoff with acceptable combination of sensitivity and specificity for most AABs in adult patients [14,15].

Taken together, there are histomorphologic as well as pathophysiological similarities between COVID-19-associated ARDS and lung manifestations of CTDs. We therefore hypothesize that a dysregulated immune response upon SARS-CoV-2 infection might reflect acute exacerbation of connective tissue disease-associated interstitial lung disease (CTD-ILD) and screened SARS-CoV-2-positive patients with comprehensive clinical follow-up as well as radiological and histopathological assessment for serological markers of autoimmunity.

## PATIENTS, MATERIAL AND METHODS

In the present study, we included all consecutive patients with positive SARS-CoV-2-RT-PCR (mucosal swab, pharyngeal or bronchoalveolar lavage) admitted to the Bundeswehrkrankenhaus Ulm in March and April 2020 after obtaining informed consent. Cases without confirmed SARS-CoV-2 infection as well as cases without/ weak symptoms and no need for hospitalization were excluded. A timeline of the complete study cohort is shown in **Fig. 1**; complete clinical data for ICU- and non-ICU patients is summarized in **Table 1**.

**Table 1.**
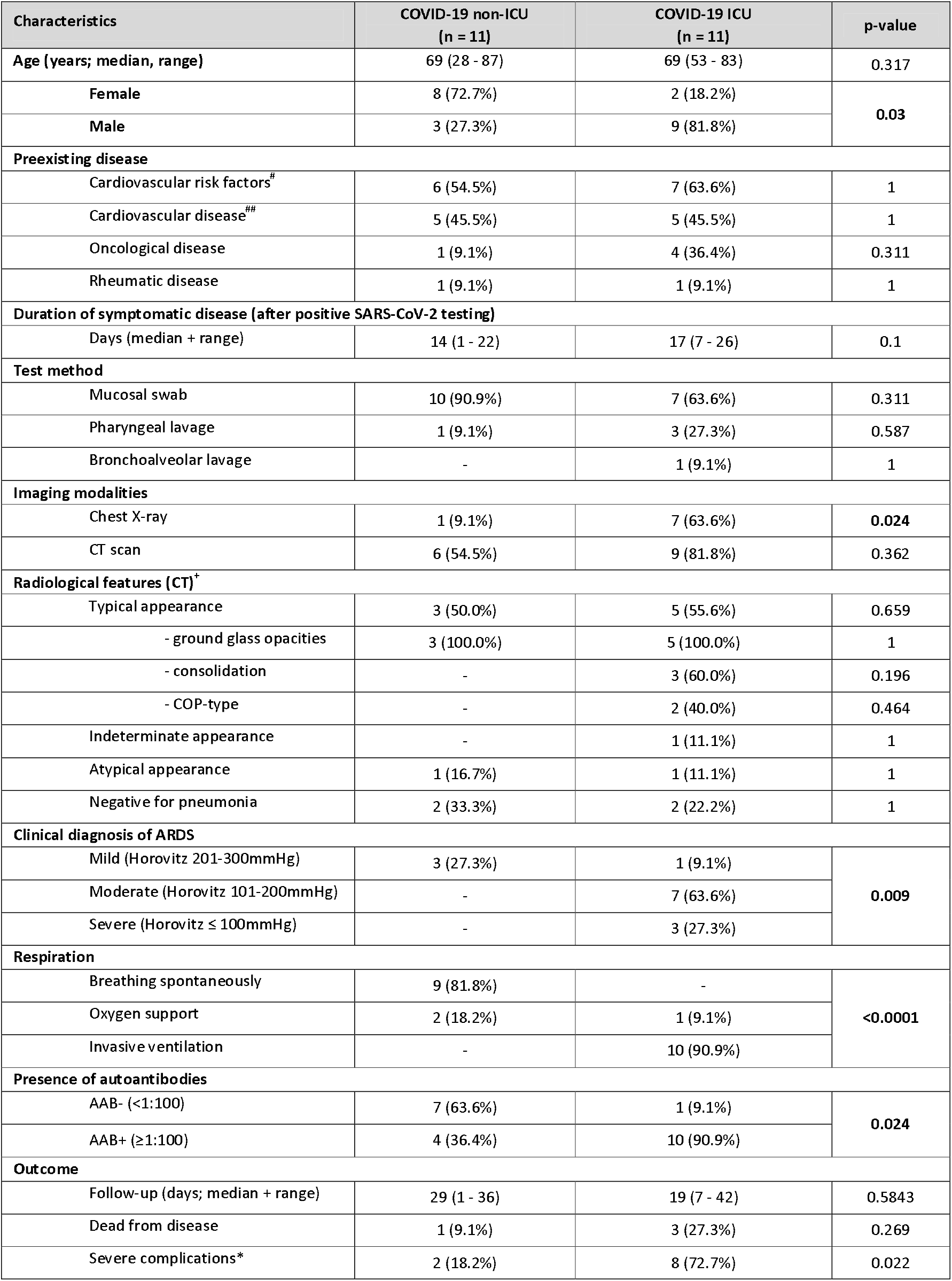
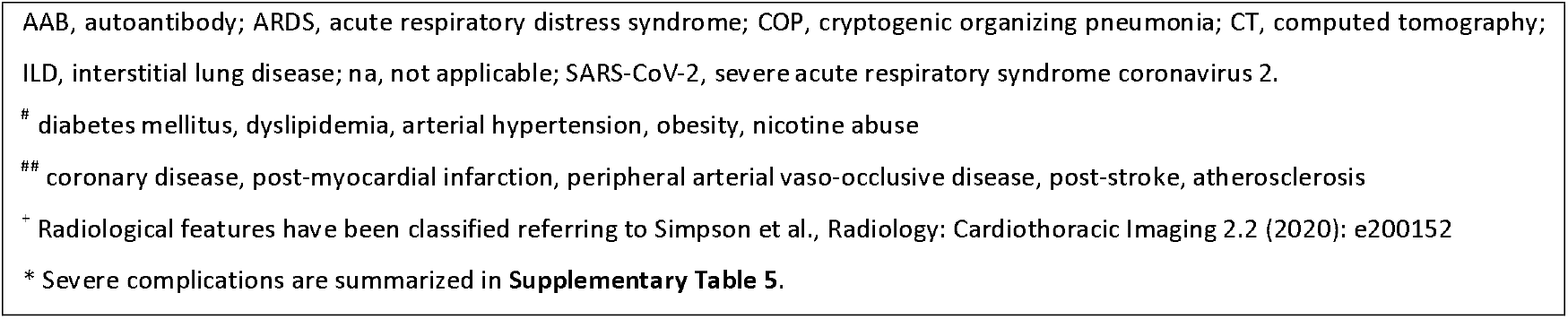
Baseline clinical characteristics of COVID-19 patients included in the study.

| Characteristics | COVID-19 non-ICU<br>(n = 11) | COVID-19 ICU<br>(n = 11) | p-value |
| --- | --- | --- | --- |
| Age (years; median, range) | 69 (28 - 87) | 69 (53 - 83) | 0.317 |
| Female | 8 (72.7%) | 2 (18.2%) | 0.03 |
| Male | 3 (27.3%) | 9 (81.8%) |  |
| Preexisting disease |  |  |  |
| Cardiovascular risk factors<sup>#</sup> | 6 (54.5%) | 7 (63.6%) | 1 |
| Cardiovascular disease<sup>##</sup> | 5 (45.5%) | 5 (45.5%) | 1 |
| Oncological disease | 1 (9.1%) | 4 (36.4%) | 0.311 |
| Rheumatic disease | 1 (9.1%) | 1 (9.1%) | 1 |
| Duration of symptomatic disease (after positive SARS-CoV-2 testing) |  |  |  |
| Days (median + range) | 14 (1 - 22) | 17 (7 - 26) | 0.1 |
| Test method |  |  |  |
| Mucosal swab | 10 (90.9%) | 7 (63.6%) | 0.311 |
| Pharyngeal lavage | 1 (9.1%) | 3 (27.3%) | 0.587 |
| Bronchoalveolar lavage | - | 1 (9.1%) | 1 |
| Imaging modalities |  |  |  |
| Chest X-ray | 1 (9.1%) | 7 (63.6%) | 0.024 |
| CT scan | 6 (54.5%) | 9 (81.8%) | 0.362 |
| Radiological features (CT)<sup>†</sup> |  |  |  |
| Typical appearance | 3 (50.0%) | 5 (55.6%) | 0.659 |
| - ground glass opacities | 3 (100.0%) | 5 (100.0%) | 1 |
| - consolidation | - | 3 (60.0%) | 0.196 |
| - COP-type | - | 2 (40.0%) | 0.464 |
| Indeterminate appearance | - | 1 (11.1%) | 1 |
| Atypical appearance | 1 (16.7%) | 1 (11.1%) | 1 |
| Negative for pneumonia | 2 (33.3%) | 2 (22.2%) | 1 |
| Clinical diagnosis of ARDS |  |  |  |
| Mild (Horovitz 201-300mmHg) | 3 (27.3%) | 1 (9.1%) | 0.009 |
| Moderate (Horovitz 101-200mmHg) | - | 7 (63.6%) |  |
| Severe (Horovitz ≤ 100mmHg) | - | 3 (27.3%) |  |
| Respiration |  |  |  |
| Breathing spontaneously | 9 (81.8%) | - | <0.0001 |
| Oxygen support | 2 (18.2%) | 1 (9.1%) |  |
| Invasive ventilation | - | 10 (90.9%) |  |
| Presence of autoantibodies |  |  |  |
| AAB- (<1:100) | 7 (63.6%) | 1 (9.1%) | 0.024 |
| AAB+ (≥1:100) | 4 (36.4%) | 10 (90.9%) |  |
| Outcome |  |  |  |
| Follow-up (days; median + range) | 29 (1 - 36) | 19 (7 - 42) | 0.5843 |
| Dead from disease | 1 (9.1%) | 3 (27.3%) | 0.269 |
| Severe complications* | 2 (18.2%) | 8 (72.7%) | 0.022 |

AAB, autoantibody; ARDS, acute respiratory distress syndrome; COP, cryptogenic organizing pneumonia; CT, computed tomography; ILD, interstitial lung disease; na, not applicable; SARS-CoV-2, severe acute respiratory syndrome coronavirus 2.

**Fig. 1.**
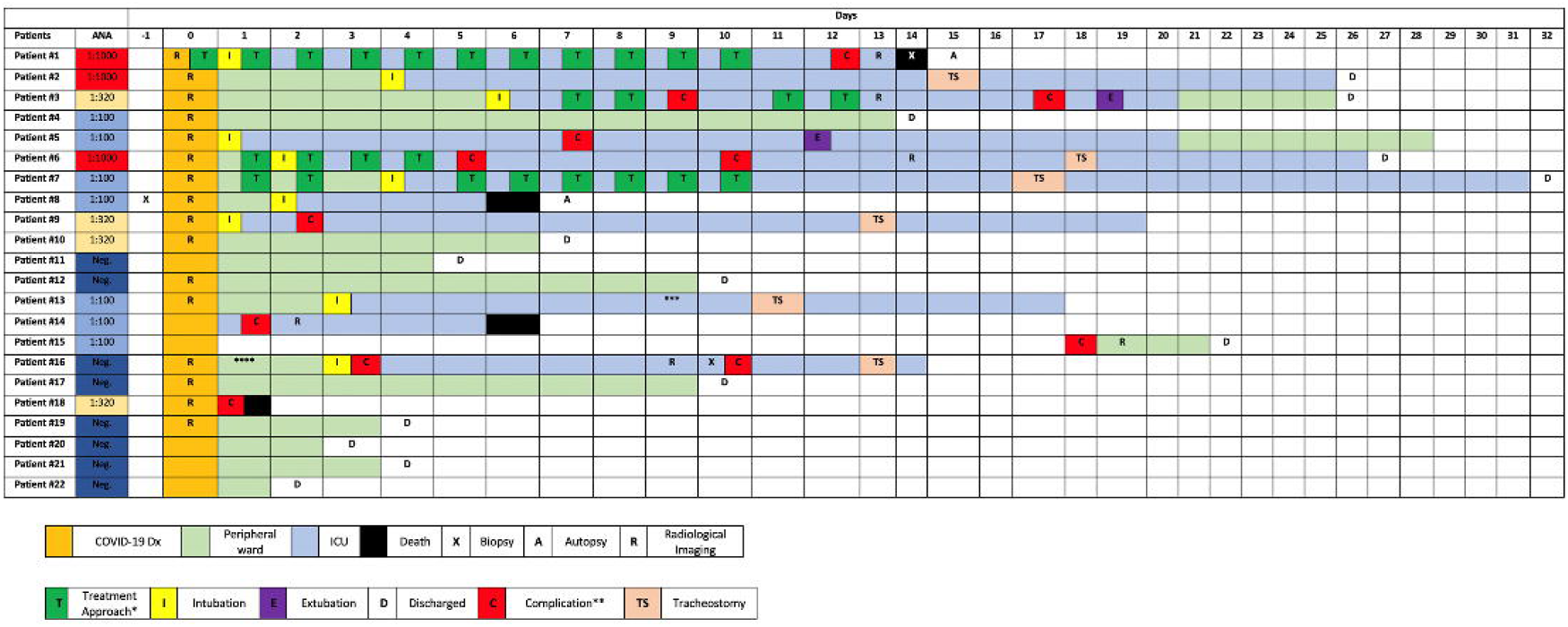
Timeline of the complete study cohort. *, treatment modalities are summarized in **Supplementary Table 1**; ** case-related complications are summarized in **Supplementary Table 5**; *** patient was transferred to another institution because vv-ECMO was required; **** patient w/ chronic lymphocytic leukemia (CLL) and antibody deficiency syndrome. Rheumatoid factor was positive in this case.

### Clinical characteristics

We collected clinical information from electronic patient files. Data collection included chronological sequence of disease-related events, preexisting comorbidities, imaging, treatment approaches (Supplementary Table 1) and clinical follow-up. The ‘’Berlin definition’’ was used to categorize ARDS [16]. The Horovitz quotient (PaO_2_/FiO_2_) was assessed in all ARDS cases based on arterial blood gas analysis.

### ICU treatment

During ICU treatment, ventilation parameters, duration of invasive ventilation, catecholamine support, prone positioning, the Murray lung injury score and the need of additional dialysis was assessed[17] (**Table 2**). Veno-venous extracorporeal membrane oxygenation (vv-ECMO) was not available at our institution. A profitable trial of prone positioning was defined by an increasing Horovitz quotient of 30mmHg or more. One entire trial covered 16 hours of sustained prone positioning.

**Table 2.**
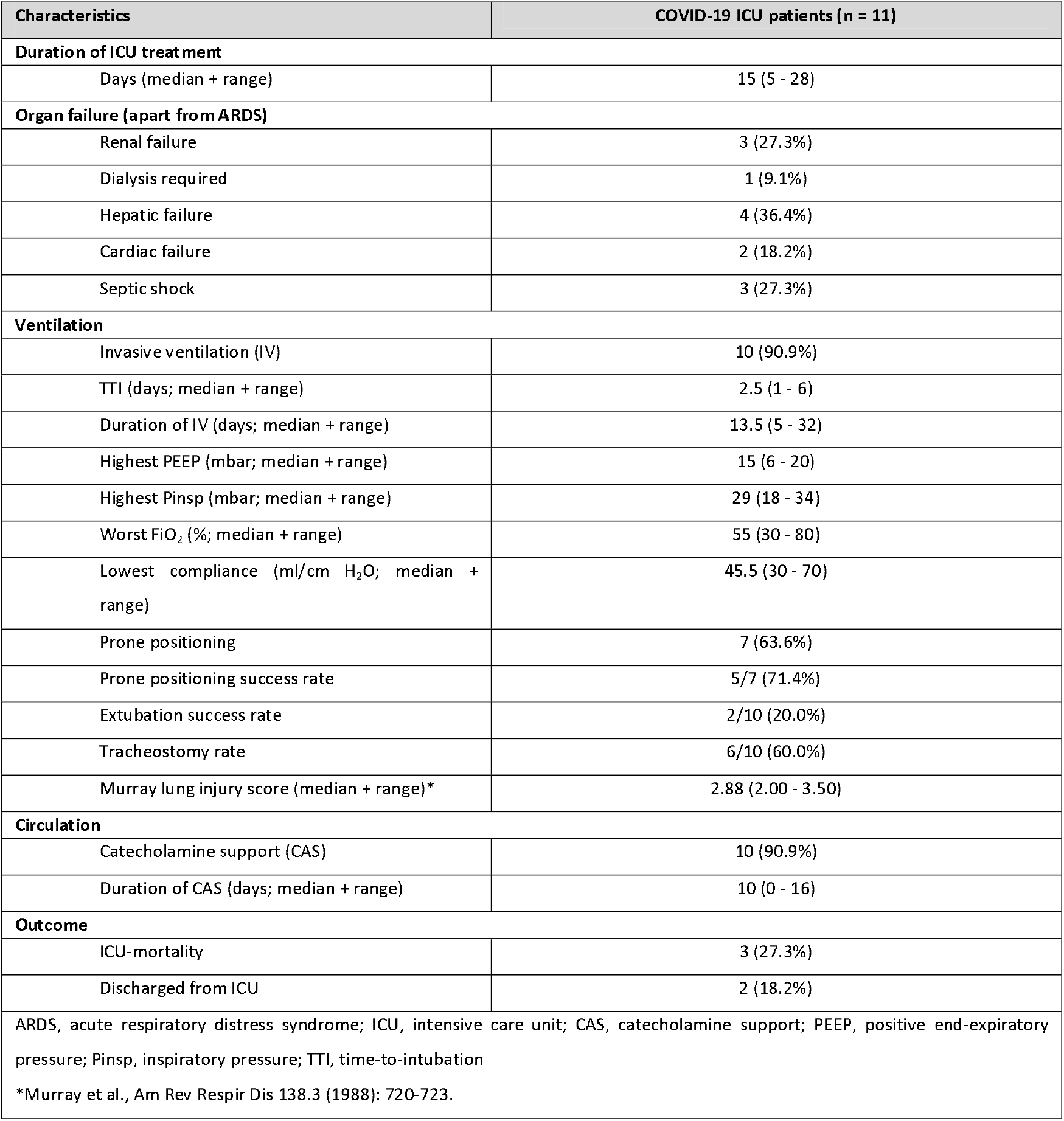
Intensive care treatment modalities.

### Serology/Laboratory values

Laboratory values included possible predictors of poor outcome in COVID-19 patients such as lymphopenia, hyperfibrinogenemia, elevated serum levels of D-dimers, ferritin, lactate dehydrogenase (LDH) and bilirubin. We also assessed troponin-T levels as a marker for cardiac events and infection-associated parameters (neutrophils, interleukin-6 (IL-6), C-reactive protein (CRP) and procalcitonin (PCT)). Cut-off values for these parameters are summarized in **Supplementary Table 2**. Furthermore, the patients were screened for autoantibody titers (summarized in **Supplementary Table 3**). Given the fact that a definite cutoff for AAB titers has so far not been, only complete absence of an AAB titer was defined as a negative result [14,15].

**Table 3.** AAB positivity and clinical characteristics of COVID-19 patients.

| Characteristics | AAB- (<1:100)<br>(n = 8) | AAB+ (≥1:100)<br>(n = 14) | p-value |
| --- | --- | --- | --- |
| Age (years; median, range) | 57 (30-88) | 71 (28-84) | 0.24 |
| Sex |  |  |  |
| Female | 5 (71.4%) | 5 (33.3%) | 0.172 |
| Male | 2 (28.6%) | 10 (66.7%) |  |
| Clinical diagnosis of ARDS |  |  |  |
| Mild (Horovitz 201-300mmHg) | 1 (12.5%) | 3 (21.4%) | 0.829 |
| Moderate (Horovitz 101-200mmHg) | 1 (12.5%) | 5 (35.7%) |  |
| Severe (Horovitz ≤ 100mmHg) | - | 3 (21.4%) |  |
| Respiration |  |  |  |
| Breathing spontaneously | 6 (75.0%) | 3 (21.4%) | 0.037 |
| Oxygen support | 1 (12.5%) | 2 (14.3%) |  |
| Invasive ventilation (IV) | 1 (12.5%)* | 9 (64.3%) |  |
| Duration of IV (days; median, range) | 12 * | 14 (5 - 32) | na |
| Radiological features (CT)<sup>†</sup> |  |  |  |
| Typical appearance | 2 (50.0%) | 6 (54.5%) | 1 |
| Indeterminate/ Atypical/ Negative | 2 (50.0%) | 5 (45.5%) |  |
| Laboratory values (median, range) |  |  |  |
| LDH (U/l) | 194 (167 - 248) | 365 (254 - 754) | 0.003 |
| CRP (mg/dl) | 0.5 (0.1 - 25.5) | 11.7 (1.0 - 42.1) | 0.075 |
| IL-6 (pg/ml) | 16 (2 - 198) | 213 (2 - 2205) | 0.125 |
| Outcome |  |  |  |
| Follow-up (days; median + range) | 25 (8 - 36) | 25.5 (1 - 42) | 0.744 |
| Dead from disease | 0 | 4 (28.6%) | 0.254 |
| Severe complications** | 1 (12.5%) | 9 (64.3%) | 0.031 |
| AAB, autoantibody; ARDS, acute respiratory distress syndrome; CRP, C-reactive protein; ICU, intensive care unit; IL-6, interleukin 6; na, not applicable. |  |  |  |
| *Pat w/ chronic lymphatic leukemia and antibody deficiency syndrome but rheumatoid factor (RF)-positive. |  |  |  |
| ** Severe complications are summarized in **Supplementary Table 5**. |  |  |  |
| † Radiological features have been classified referring to Simpson et al., Radiology: Cardiothoracic Imaging 2.2 (2020): e200152 |  |  |  |

### Imaging

Imaging for COVID 19 was performed on a Somatom Force Scanner (Dual Source Scanner 2*192 slices, Siemens, Erlangen, Germany) in accordance to the guidelines of the German Radiological Society and our hospital’s COVID-19 guidelines, using low-dose CT (computed tomography) with high-pitch technology[18]. The following parameters were used: Tube voltage: 100kV with tin filtering, tube current: 96 mAs with tube current modulation. In 2 cases the examination was performed as a non-contrast enhanced full-dose protocol because of suspected ILD, in one case as a contrast enhanced CT to exclude for pulmonary embolism. The X-ray examinations were performed at the respective wards as bed-side X-ray examinations (Mobilett Mira Max, Siemens, Erlangen, Germany) as a single anterior-posterior view. The CT images were evaluated according to the Expert Consensus Statement of the RSNA and classified as typical, indeterminate, atypical and negative appearance for COVID[18,19].

### Histology and immunohistochemistry

Lung tissue specimens were obtained as transbronchial biopsies in 3 cases, one of which had been tested SARS-CoV-2 negative in previous mucosal swab. In two patients, limited autopsies were performed after death and lung, heart and liver tissue was sampled extensively. Specimens were stained with haematoxylin-eosin (HE), Elastica-van Gieson (EvG) and Masson-Goldner (MG). Furthermore, immunohistochemistry for CD3, CD68, CMV and EBV was performed using prediluted antibodies on a VENTANA benchmark autostainer (Roche Tissue Diagnostics, Mannheim, Germany) following routine protocols.

### Ethics statement

Patients or their relatives had given written informed consent to routine diagnostic procedures (serology, bronchoscopy, radiology) as well as (partial) autopsy, respectively, as well as to the scientific use of data and tissue samples in the present study. This project was approved by the local ethics committee of the University of Ulm (ref. no. 129-20) and conducted in accordance with the Declaration of Helsinki.

### Statistical methods

Descriptive statistical methods were used to summarize the data. Medians and interquartile ranges were used to announce results. Counts and percentages were employed to represent categorial variables. Student’s t-test was used for the comparison of continuous variables, while Chi-Square-Test/Fisher’s test was used for categorical varoiables. All statistical analyses were conducted using GraphPad PRISM 6 (GraphPad Software Inc., San Diego, CA, USA). A p-value<0.05 was regarded as statistically significant.

## RESULTS

### Baseline clinical characteristics

Baseline clinical characteristics of SARS-CoV-2 infected patients included in the current study are briefly summarized in **Table 1**. The median follow-up period in the present study cohort was 25.5 days (range 1-42 days, 25% percentile 21.0 days; 75% percentile 30.0 days). Median age at initial diagnosis was 69.0 years (range, 28 - 88 years). A majority of patients was male (12/22 cases; 54.5%). Mucosal swabs detected 81.8% of SARS-CoV-2 infections while pharyngeal lavage revealed 13.6% of positive test results. 4.5% of cases were tested positive by RT-PCR of bronchoalveolar lavage. Preexisting oncological or immunosuppressive diseases were present in 7/22 cases (31.8%). The most frequent preexisting illness was cardiovascular disease (10/22 cases; 45.5%). Spontaneous breathing was sufficient in 9/22 patients, whereas oxygen supply via nasal cannula was required in 3/22 cases. 13/22 cases (59.1%) presented with or developed ARDS according to the ‘’Berlin-definition’’[16].

Antiviral treatment/inhibition of autophagy was applied in 4/22 (18.2%) patients. Median duration of these approaches was 3 days (range 2-10 days). The toxicity profile was moderate and predominantly nephrotoxic in nature with 2 cases of acute kidney injury Kidney Disease Improving Global Outcome (KDIGO) stage III. Other toxicities included drug-related exanthema in one patient. Application of hydroxychloroquine (HCQ) was completed when COVID-19 patients from other hospitals were transferred to our institution. Antiviral treatment approaches were discontinued immediately upon transfer. Treatment approaches are summarized in **Supplementary Table 1**.

Bacterial superinfection was suspected in 10/22 (45.5%) cases by clinical and laboratory presentation and imaging. Antibiotic treatment approaches are summarized in **Supplementary Table 4**. The mortality related to SARS-CoV-2 infection in current study population was 18.2% (4/22 patients). There was a high rate of complications (**Supplementary Table 5**). One patient had to be transferred to another institution because vv-ECMO was required. After a median follow-up period of 29 (non-ICU patients) and 19 days (ICU) patients, 4 patients (18.2%) had died from disease.

### Clinical course and intensive care (ICU) treatment

In total, intensive care treatment was required for 11/22 (50.0%) COVID-19 patients. While more ICU patients were of male sex (p=0.03), there were no significant differences in age or preexisting comorbidities between ICU and non-ICU patients. ICU treatment modalities and ventilation parameters are summarized in **Table 2**. Invasive ventilation was necessary in 10 cases. Median duration of invasive ventilation was 13.5 days (range 5 - 32 days). The majority of intubated patients underwent tracheostomy (6/10 cases; 60.0%), whereas extubation was successful in 2 patients (20.0%) until the end of follow-up. Arterial blood gas analysis revealed moderate ARDS in the majority of COVID-19 patients (7/13 cases; 53.8%) while severe ARDS was found in 3 patients (23.1%) and mild ARDS in 3 patients (23.1%) by the Horovitz quotient[16]. Apparently, ICU treatment was significantly associated with severity of clinical ARDS (p=0.009) and assisted/invasive ventilation (p<0.0001; **Table 1**). Prone positioning was performed in 7 cases, which was successful in five patients (71.4%). The Murray lung injury score was calculated for all patients that underwent invasive ventilation. This score revealed severe ARDS in 8/10 cases (80.0%)[17]. Median duration of catecholamine support was 10 days in COVID-19 patients admitted to intensive care unit.

### Laboratory findings/ serology

Characteristic laboratory constellations in COVID-19 cases were associated with elevated serum levels of lactate dehydrogenase (66.7%; median 297.5 U/l; range 167 - 754 U/l) and bilirubin (52.4%; median 0.69 mg/dl; range 0.23 - 5.41 mg/dl) **(Supplementary Table 2**). There was high procoagulant activity shown by elevated levels of fibrinogen (100.0%; median 5.16 g/l; range 2.74 - 9.00 g/l) and D-dimers (71.4%; median 1.48 mg/l; range 0.31 - 30.00 mg/l). Elevated levels of CRP (76.2%), IL-6 (85.7%) and TNF-alpha (100.0%) were found in the majority of cases. Lymphopenia was present in 16/22 cases (72.7%).

Screening for autoantibodies revealed positive ANA titers in the majority of cases with moderate or severe ARDS and in 90.9 % (10/11) ICU cases compared to 36.4% (4/11) non-ICU cases (p=0.024). The distribution and type of AAB titers among the ICU/non-ICU patients is shown in **Fig. 2**. The single female ICU patient (#22) without detectable autoantibodies presented with CLL-associated antibody deficiency syndrome but with a history of rheumatoid factor-positive rheumatoid arhritis. One patient with detectable autoantibodies (#18, ANA 1:320) died from severe ARDS and pulmonary embolism before he could be transferred to the ICU. Presence of AAB titers was associated with higher age and male sex, although not significant. AAB positivity, however, was associated with a necessity of assisted/invasive ventilation (p=0.037) and occurrence of severe complications (p=0.31). Distribution of typical or atypical patterns in CT imaging were not different between patients with and without AABs. There was no significant association between AAB status and disease-specific survival in the investigated cohort.

**Fig. 2.**
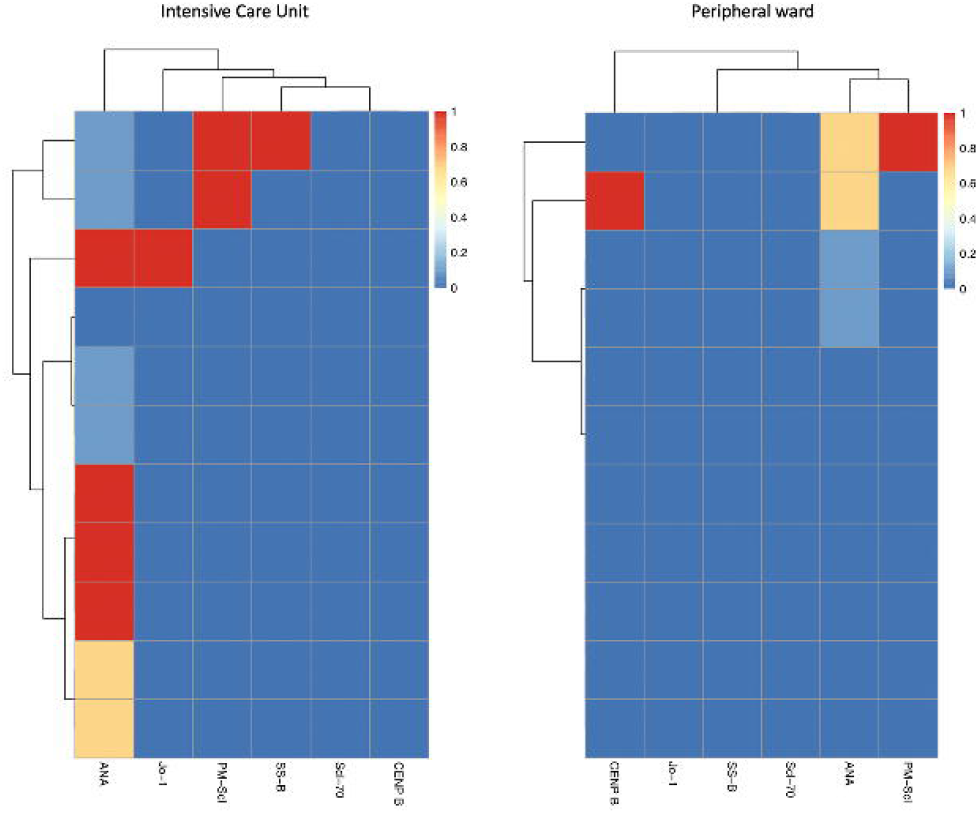
Heatmap showing the distribution and subtype of autoantibody titers in the group of ICU (left) and non-ICU patients (right).

### Imaging and Histopathology

“Typical” radiologic COVID-19 patterns were found in 53,3% of patients (ICU: 55,6%/non-ICU: 50%). These included ground glass opacities (all “typical” cases), consolidation and COP-like pattern (Fig. 3). Atypical/negative patterns were found in 44,4% of ICU and 50% of non-ICU patients. Bronchoscopy with transbronchial biopsy (TBB) was performed in three patients (cases #1, #8 and #16) before (#8) and after (#1, 16) an established diagnosis of COVID-19, respectively (**Fig. 1**). From two of these patients (#1, 8), additional tissue samples were obtained during a limited autopsy procedure. In all samples, we observed reactive pneumocyte changes (“Napoleon hat sign”) consistent with viral infection (**Fig. 4**). However, there was a marked variance in the histologic appearance between different patients, between TBB and autopsy samples from the same patient and between autopsy samples from different regions of the lung. In addition to hyaline membrane formation consistent with diffuse alveolar damage (DAD), there was also septal thickening as well as alveolar fibrinous plug formation with partial fibromyxoid change, reminiscent of acute fibrinous organizing pneumonia (AFOP)(#1, **Fig. 4 A**). In Patient #8, there was predominantly interstitial organization without evidence of DAD (**Fig. 4 B**).

**Fig. 3.**
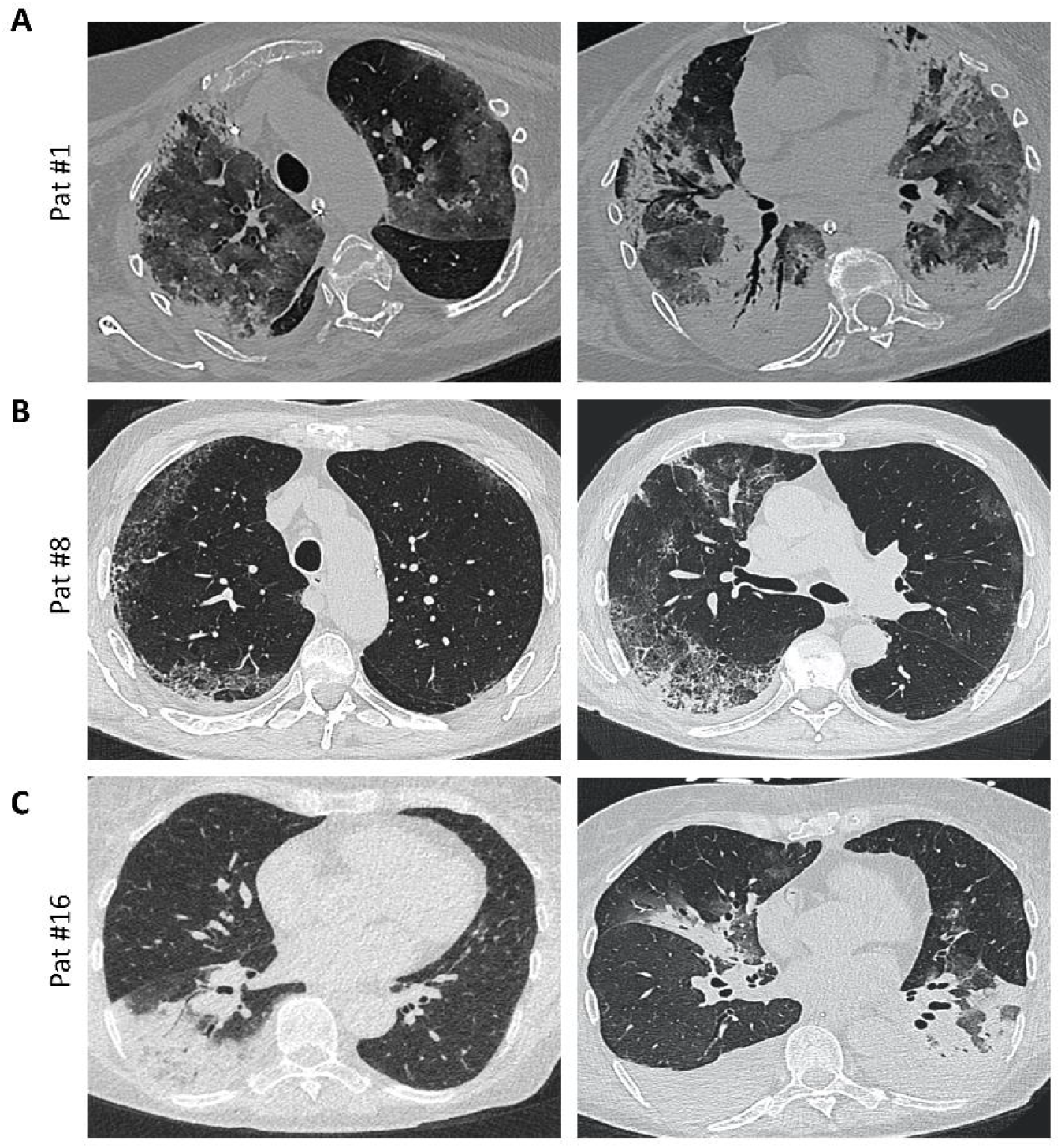
Imaging. A, Axial thin-section unenhanced CT scan of a 69-year-old woman (patient #1, ANA 1:1000; see **Fig. 4 A** for histology) 12 days after admission with later-stage disease. Mixed image with diffuse ground-glass opacities (GGO) and subpleural and peribronchial distributed consolidations with positive aerobrochogram and resembeling an organizing pneumonia. Additionally, anteriorly accentuated irregular subpleural consolidations with partially left-out subpleural space. **B**, Axial thin- section unenhanced CT scan of a 80 year-old male (patient #8; ANA 1:100; see **Fig. 4 B** for histology), imaged for suspected interstitial lung disease (ILD). Images show right-sided dominant fibrotic changes in the periphery, with partially sparing of the subpleural space, resembling a NSIP-like pattern. Minimal ground-glass-opacities (GGO) in the left subpleural space. **C**, Axial thin-section unenhanced CT scans of a 66 year- old female (patient #16, ANA negative, but history of RF+ RA). The left image was obtained on the day of hospitalization with a typical sign of a lobar pneumonia of the right lower lobe. The right image (day 10 after admission) shows bilateral ground glass opacities and consolidations, mainly on the left lower lobe and the middle lobe. The left lower lobe is once again properly aerated. Additionally, bilateral pleural effusions are detectable.

**Fig. 4.**
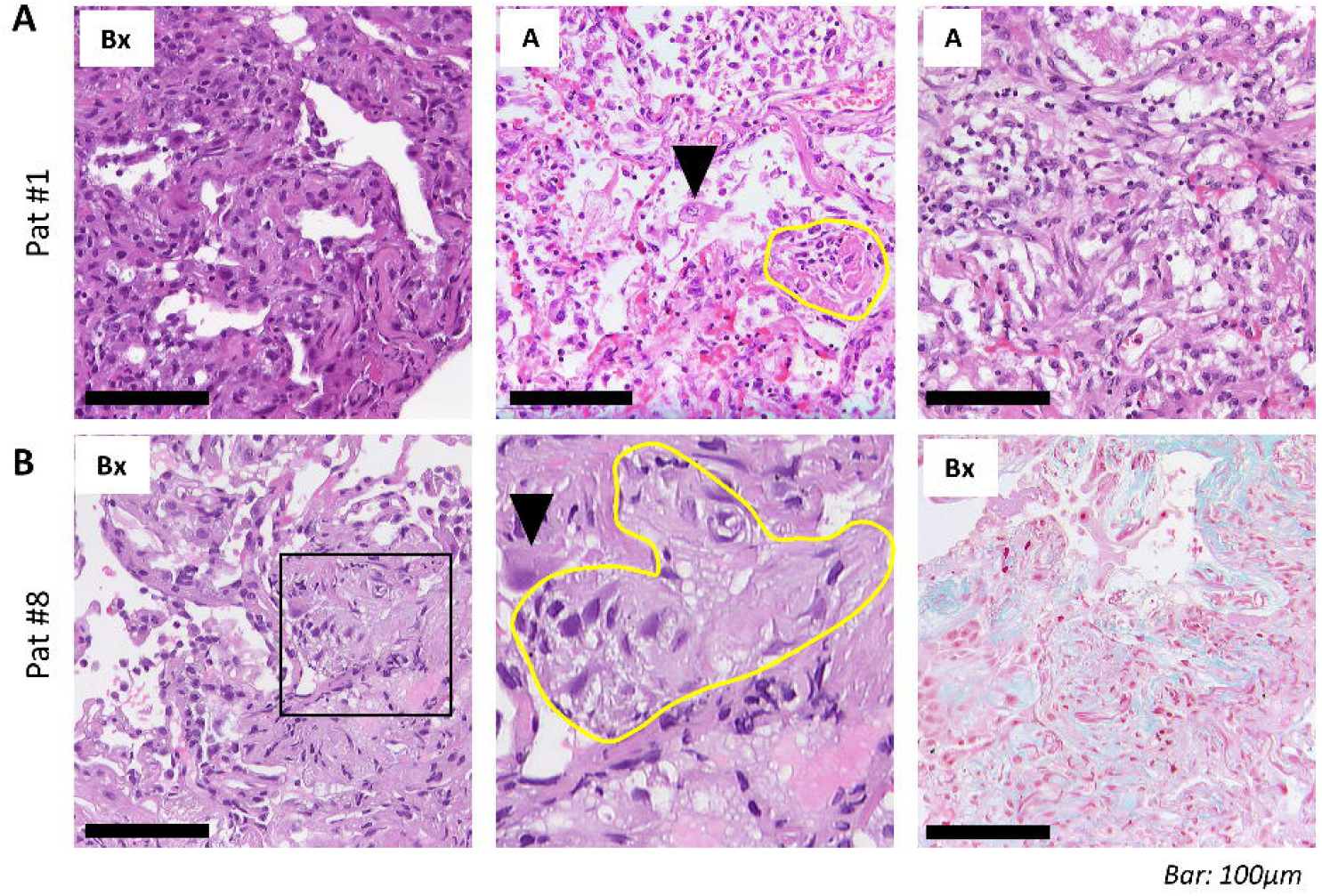
Histopathological assessment. A, Transbronchial biopsies (Bx, left) in a 69-year-old woman (patient #1, ANA 1:1000; see **Fig. 3 A** for imaging) 13 days after admission shows septal thickening without fibrinous exudate. Tissue samples from the autopsy (A, center) of the same patient with reactive pneumocyte changes (“Napoleon hat sign”, arrowhead), ball-like fibrin (yellow circle) and alveoli with plug-like fibromyxoid organization (right). **B**, Transbronchial biopsies in an 80-year-old man (patient #8, ANA 1:100; see **Fig. 3 B** for imaging) show reactive pneumocyte changes (arrowhead) and interstitial fibroblastic foci (yellow circle). Masson-Goldner stain (right) highlights subtle interstitial fibrosis (bright green). *Scale bar, 100µm*.

## Discussion

In the present study, we investigated the prevalence of autoantibodies in a cohort of 22 consecutive patients with confirmed SARS-CoV-2 infection (COVID-19) and possible correlations with clinical, radiologic and histopathological findings. We show that the presence of AABs is significantly associated with the necessity for ICU treatment and invasive ventilation as well as occurrence of severe complications in these patients; noteworthy, every ICU patient in the present study had detectable autoantibodies and/or history of autoimmune disease. With respect to baseline clinical characteristics, the investigated cohort is comparable to previous reports [2,3]. Notably, in our study, ICU patients/patients with severe clinical course or patients with AABs were not significantly older or had more preexisting comorbidities compared to patients with a mild clinical course of the disease or AAB-negative patients, respectively. However, we could confirm a high rate of lymphopenia as well as elevated LDH levels in COVID-19 patients, consistent with data from the literature [2]. While we can confirm from our cohort that hyperfibrinogenemia and elevated D-dimers are frequent in COVID- 19 patients, only two patients presented with thromboembolic events. Given the fact that only hospitalized patients were included, it is not surprising that the mortality rate (18.2%) was higher compared to the general population.

The imaging and histopathological data presented here and in previous studies show that the presentation of COVID-19 in the lung can be quite heterogeneous [5,20,21]. The overlapping features of the observed changes including diffuse alveolar damage, fibromyxoid plugging and interstitial fibrosis are reminiscent of features associated with exacerbation of CTD [22,23], in line with our main finding of AABs in patients with a severe course of COVID-19. However, it is unclear whether these AAB were already present in predisposed patients prior to infection or if transient appearance of AABs is induced by the virus, as has been described for different types of viral infections [24]. A possible epiphenomenon has therefore to be discussed. However, it is unlikely that the transient development of a non-specific ANA-IgG titer as a “price to pay” during the fight against SARS-CoV-2 infection would occur and reach titers of up to 1:1000 in such limited time [25]. In addition, only few of the non-ICU patients showed AAB despite a long phase of virus colonization in these cases. However, we cannot exclude that a certain COVID-19-specific immunogenic stimulus contributes to the development of AABs in a subgroup of patients, which would still offer the possibility to stratify patients according to their individual risk at an early point during the course of the disease. In light of the results presented here, there are interesting parallels between the reported epidemiology of severe COVID-19 and the presence of autoantibodies in the general population. Autoantibody titers above 1:80 and 1:160 can be detected in 13.3% and 5% of otherwise healthy individuals [15], reflecting reported proportions of severe (14%) and critical (5%) course of COVID-19 [26]. Preliminary reports from the U.S. suggest that the COVID-19-associated death rate among African Americans is significantly higher compared to the general population [27], while at the same time ANA titers in African Americans exceed those of Americans with another ethnic background [28]. From our perspective, higher ANA titers in African Americans might therefore contribute to the development of severe clinical courses of COVID-19.

There is an ongoing debate with regard to a possible dysregulation of the immune system by SARS-CoV-2, and it has been discussed whether anti-inflammatory drugs might be beneficial to prevent potentially harmful hyperinflammation [29]. Previous studies have demonstrated that coronaviruses activate toll-like receptors (TLRs) on antigen-presenting cells (APCs), followed by a release of IFNα and functional exhaustion of cytotoxic T-lymphocytes (CTL) [10,11]. It has been shown that such decrease in natural killer (NK) and CD8^+^; cytotoxic T-cells paralleled by a relative increase of NKG2A+ NK- and CD8+ T-cells correlates with disease progression in COVID-19 patients [1]. NKG2A was identified as a potential immunotherapeutic target that may prevent from functional exhaustion of CTLs and contributes to elimination of cells infected by SARS-CoV-2 while several antiviral treatment approaches did not improve clinical outcome significantly in COVID-19 patients with moderate or severe ARDS [30]. In line with that, upregulation of NKG2A is a marker for functional exhaustion of CTLs in cancer patients and patients suffering from chronic viral infections. These mechanisms show high similarity to the development and progression of connective tissue disease (CTD), such as systemic Lupus erythematodes (SLE), Sjögren’s, or systemic sclerosis (SS)[12]. For example, NK cells from SLE patients with high disease activity showed increased expression of NKG2A while there was functional exhaustion of CTLs [13]. Our hypothesis of SARS-CoV-2-induced immune dysregulation closely correlates with results from the Wuhan cohort reported by Wu et al., in which methylprednisolone treatment was associated with a more favourable outcome among the patients who had already developed ARDS [2]. It would be interesting to evaluate if patients with SLE-like AAB pattern (ANA, anti-ds-DNA) profit from chloroquine, while patients with an SSc-like AAB pattern (anti-Scl, anti-centromer) might respond to cyclophosphamide. However, the correct timing and dose for any immunosuppressive treatment approach in response to a viral infection remains unclear.

To our best knowledge, we are the first to show relevant AAB titers in patients with a severe course of COVID-19. Together with the heterogeneity in radiologic and histopathologic findings we present here, our results point towards a possible dysregulation of the immune response upon SARS-CoV-2 infection that might be comparable to lung involvement in acute exacerbation of autoimmune disease. The early detection of autoantibodies might identify patients who profit from immunosuppressive therapy to prevent the development of respiratory failure in COVID-19.

## Data Availability

All original data can be obtained from the authors.

## Author contributions

Study concept: DG and KS. Data collection: HMW, DG, MH, VU, JS, KS. Sample collection: DG, VU. Statistical analysis: HMW, NG and KS. Initial draft of manuscript: KS, JS, DG and HMW. Critical revision and approval of ?nal version: all authors.

## Acknowledgments

The authors would like to thank all patients and their families for their consent to the use of data and images in the present study. We further thank Judith Bauer, MD and Stephan Opderbeck, MD for providing clinical data. The authors are grateful for the outstanding quality of care of COVID-19 patients provided by the team of the intensive care unit (ICU) at the Bundeswehrkrankenhaus Ulm.

